# The association between medical accessibility and glycemic status in patients with type 2 diabetes mellitus: A retrospective study in Taiwan rural population

**DOI:** 10.1101/2024.10.18.24315733

**Authors:** Chong-Fong Sun, Bo-Lin Pan, Pei-Ming Wang, Kun-Siang Huang, Yueh-Chien Lu, Hsin-Yi Kung, Jui-Chin Chiang

**Affiliations:** Department of Family Medicine, Kaohsiung Chang Gung Memorial Hospital, Kaohsiung City, Taiwan; Department of Nursing, Kaohsiung Chang Gung Memorial Hospital, Kaohsiung City, Taiwan

## Abstract

**Introduction:** Type 2 diabetes mellitus represents a significant chronic health concern, with rural populations being particularly vulnerable due to geographic and socio-economic barriers to healthcare access. To date, few studies have focused on the medical accessibility of rural diabetic patients in Taiwan. This study aims to explore the association between the proximity to healthcare facilities and glycohemoglobin levels in the rural Taiwanese population.

**Methods:** We conducted a retrospective cross-sectional study in rural districts of southern Taiwan. Medical accessibility was calculated from subject’s address to medical units by Google Maps platform. Spearman’s Rank Analysis was performed for the correlation between distance from patient’s address to medical units and glycohemoglobin level. Logistic regression was applied to identify factors associated with glycemic control.

**Results:** In total, 117 patients with type 2 diabetes mellitus were enrolled (mean glycohemoglobin: 8.17±1.92%). Glycohemoglobin level was modestly and positively associated with distance to clinic (Rho: 0.252) and hospital (Rho: 0.241) respectively. In logistic regression analysis, good glycemic control (glycohemoglobin less than 7%) was associated with covariates as not-working status, the address district near downtown, fasting plasma glucose equal to or less than 130 mg/dL, and normal high-density lipoprotein cholesterol level.

**Conclusion:** Our study reveals that accessibility to healthcare facility is modestly correlated with long-term glycemic status in Taiwan rural diabetic population. Besides, we also identified several protective factors associated with chronic glycemic status. These findings may pave the way for improvement of policy and healthcare strategies in rural diabetic care.

## Introduction

Type 2 Diabetes Mellitus (T2DM) is the predominant form of diabetes worldwide, comprising approximately 90% of all cases. According to the 10th edition of the International Diabetes Federation Atlas, the global diabetic population is projected to reach 783.2 million by 2045, marking an increase of nearly 50% from the estimates made in 2019[1]. The prevalence of diabetes mellitus in Taiwan aligns with this global upward trend. Data from a retrospective longitudinal cohort study using the Taiwan National Health Insurance Research Database indicated a 66% increase in the diabetic population from 2005 to 2014[2]. T2DM is defined by chronic hyperglycemia, which can silently damage multiple organs. If not properly managed, T2DM can lead to severe complications such as stroke, blindness, and amputation of lower extremities, thereby disabling individuals and imposing significant economic and healthcare burdens. Bommer, C. et al. highlighted that global healthcare expenditures for diabetes among individuals aged 20-79 increased from USD 232 billion in 2007 to USD 966 billion in 2021[3]. In Taiwan, the frequency of outpatient visits by T2DM patients is roughly double that of the average patient[4]. Patients with diabetic complications also tend to have more emergency room visits and hospitalizations, which result in higher and rising costs under Taiwan’s National Health Insurance program[4, 5].

Effective prevention of diabetic complications hinges on self-care and regular medical check-ups, considered as the foundation of diabetes management[1, 6]. However, it is challenging for rural T2DM patients to access continuous diabetic care due to geographical and financial barriers. Dugani, S. B. et al. reported that rural populations in the United States have a higher prevalence of T2DM and diabetic hospital mortality rates-16% and 20%, respectively-compared to urban populations[7]. Similarly, an observational study in rural China found that nearly 60% of diabetics were unaware of their health condition, and 40% of those diagnosed with T2DM were not receiving treatment[8]. These factors contribute to the higher incidence of diabetes-related complications among rural populations. In Taiwan, national database studies have shown that rural T2DM patients are more likely to develop ischemic heart disease, lower limb ulcers, stroke, congestive heart failure, end-stage renal disease, and blindness[5]. Additionally, delayed detection of diabetic neuropathy and an approximately 50% higher prevalence of lower limb amputations were reported[9, 10]. Rural T2DM patients also have a higher risk of preventable hospitalizations compared to their urban counterparts[11].

Considering the rural-urban disparities, recent studies have explored the impact of distance to healthcare facilities on diabetes management[12–16]. Nevertheless, to our knowledge, no study has specifically addressed this issue in Taiwan. This study aims to illuminate the relationship between geographic accessibility and long-term glycemic control, and to identify protective factors that promote good glycemic management in Taiwan’s rural diabetic population.

## Methods

### Study design and subjects

We conducted a retrospective, cross-sectional study in two rural districts, Jia-Sian and Shan-Lin, in southern Taiwan, spanning from January 2009 to December 2022. The study participants were selected from those using the outpatient service of the Mobile Health Care program. This public program, initiated by the Taiwanese government’s Ministry of Health and Welfare in 2002, aims to mitigate urban-rural health disparities. It mobilizes medical teams from urban medical centers to provide daily outpatient care, perform routine blood tests, and prescribe medications at public venues like temples or community centers. During the study period, a total of 3,082 individuals utilized this outpatient service. After verifying their T2DM diagnoses and excluding those with unavailable addresses, 117 subjects were finally included in this study (Fig. 1). The Chang Gung Medical Foundation’s Institutional Review Board approved this study (IRB No. 202301135B0).

**Fig 1.**
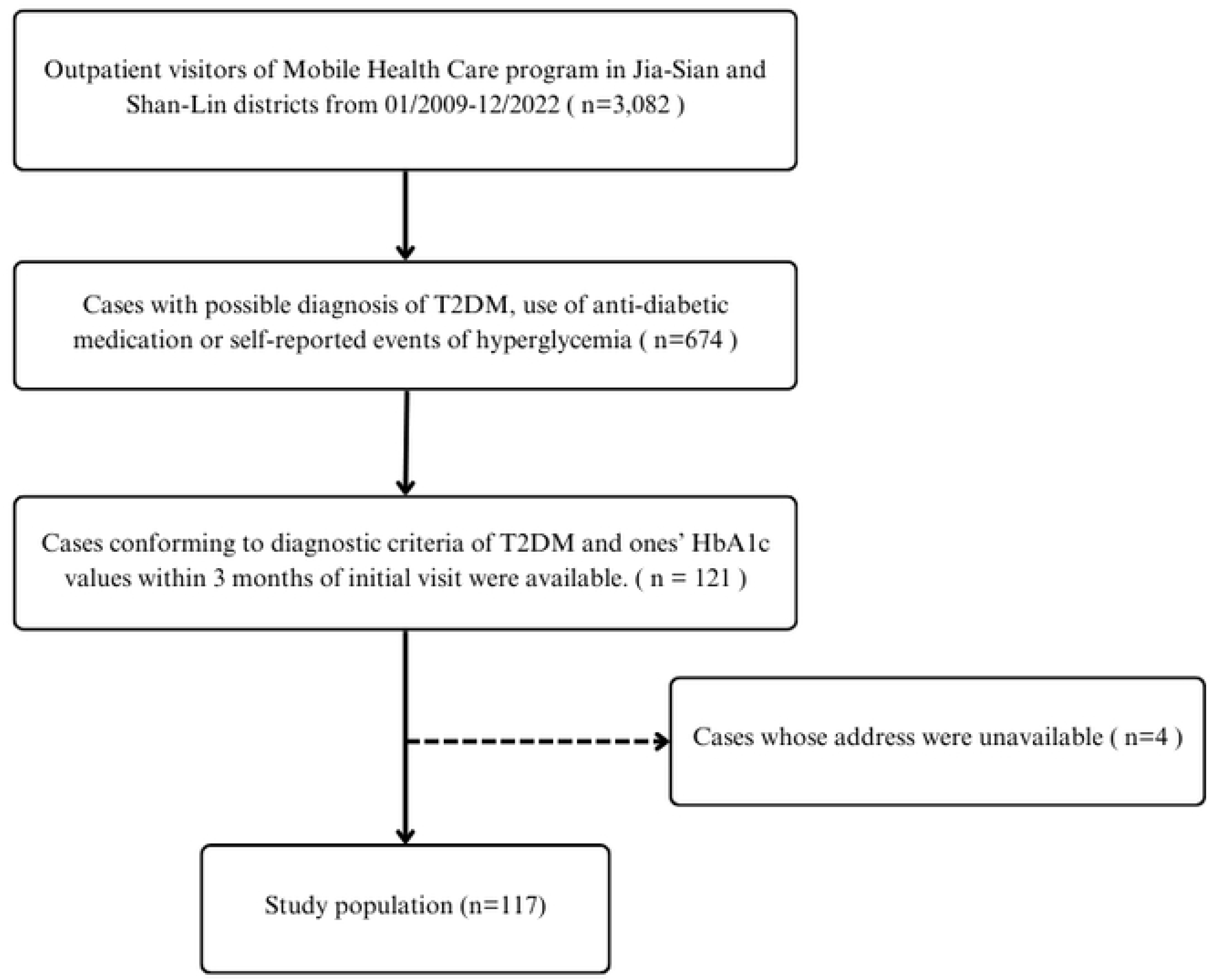
Flow chart of subject inclusion in this study. Abbreviations: T2DM, Type 2 diabetes mellitus; HbA1c, glycohemoglobin

### Data collection and laboratory methods

We started to retrospectively access patients’ data after improvement of the Chang Gung Medical Foundation’s Institutional Review Board on September 1^st^, 2023. Medical records and laboratory data for the study were extracted from the database of Kaohsiung Chang Gung Memorial Hospital, which the urban medical team belonged to. Patient profiles were compiled from the initial visit forms filled out by patients with the help of medical staff. Venous blood samples were collected from patients following a minimum of 8 hours of overnight fasting. Fasting plasma glucose level was measured by enzymatic method. Total cholesterol (TC), high-density lipoprotein cholesterol (HDL-C), and triglyceride (TG) were measured by colorimetric method. Low-density lipoprotein cholesterol (LDL-C) level was calculated by Friedewald Formula (LDL-C = TC – HDL-C – TG/5). HbA1c level was quantified through High Performance Liquid Chromatography (HPLC), and proteinuria was assessed by using a urine test that employs reflective photometry. During data collection and analysis, all subjects’ data were de-identified. The author responsible for collecting addresses and processing related data handled this personal information independently, and he did not have access to other patients’ data.

### Definition of T2DM

T2DM was defined according to diagnostic criteria outlined by Diabetes Care 2023[17]. The criteria include: (1) a patient with classic symptoms of hyperglycemia and a random plasma glucose ≥ 200 mg/dL, or (2) a patient has at least two records of fasting plasma glucose ≥ 126 mg/dL or (3) HbA1c ≥ 6.5%. Additionally, subjects who had been diagnosed with T2DM or who were taking anti-diabetic medications were also considered as having T2DM.

### Variables

In this study, we measured geographic accessibility to diabetic care by calculating the distance from a patient’s residence to the nearest medical facility, such as a hospital or clinic, using the Google Maps platform. HbA1c was employed as the surrogate of long-term glycemic status. To mitigate potential confounding effects from the medical services provided by the urban medical team of the Mobile Health Care program, we considered only HbA1c values obtained within three months before or after the patient’s initial visit. HbA1c levels were categorized into two groups: “good control,” defined as levels below 7%, and “poor control,” defined as levels equal to or exceeding 7%.

### Covariates

We categorized other variables into general data, past history, and laboratory data. General data included: (1) age, grouped as under 65 or 65 and older; (2) gender, classified as male or female; (3) employment status, either employed or not working; (4) education level, categorized as unschooled, elementary school, junior high school, or senior high school and above; (5) district, either Jia-Sian or Shan-Lin; (6) lifestyle factors, including current smoking, drinking, and betelnut use. Past history contained cardiovascular diseases, hypertension, chronic hepatitis, stroke, chronic kidney disease, malignancy, and surgical interventions. Laboratory data comprised fasting plasma glucose, lipid profiles (total cholesterol, LDL-C, HDL-C, and triglycerides), and proteinuria. Fasting plasma glucose levels of 130 mg/dL or lower, HDL-C levels of 40 mg/dL or higher in men and 50 mg/dL or higher in women, and triglyceride levels below 150 mg/dL were defined as “good control” following the recommendations from Diabetes Care 2023[17]. Total cholesterol levels below 160 mg/dL and LDL-C levels under 100 mg/dL were also considered “good control” based on the suggestion from the Diabetes Association of the Republic of China (Taiwan) [18]. Proteinuria was classified as positive (ranging from 1+ to 4+) or negative (trace and negative).

### Statistical analysis

General characteristics of subjects were presented by means and standard deviations (SD) for continuous variables and percentages for categorical variables. To explore the relationship between HbA1c levels and geographic distance to medical facilities, Spearman’s rank correlation test was employed. Chi-Square test of independence and Fisher’s exact test were used for categorical variables, depending on the expected numbers in classes. Logistic regression was utilized to estimate the odds ratio of categorical variables. All statistical analyses were conducted by using IBM SPSS software, version 22.0. All tests were two-sided, and *p*-values less than 0.05 were considered statistically significant.

## Result

### General characteristics

As presented in Table 1, the study population predominantly consisted of midlife to elderly adults, with an average age of 64.79 (±11.20) years. Approximately 60% of the participants were female. The majority of subjects had a relatively low level of education, with 73.4% attaining primary school education or less. Regarding residential districts, a higher proportion of patients (63.2%) resided in the Shan-Lin district. Among past medical histories, hypertension emerged as the most prevalent disease, affecting 76.1% of the subjects. Cardiovascular and chronic kidney diseases were the next most common conditions, occupying 27.4% and 15.4% of the participants, respectively.

**Table 1.** General characteristics.

| Characteristics | Categories | All(n%) |
| --- | --- | --- |
| Total |  | 117(100.0) |
| General data |  |  |
| Age(years) | Mean ± SD | 64.79 ± 11.20 |
| | $\geq 65$ | 54(46.2) |
| Sex | Female | 70(59.9) |
| Education level | Unschool | 39(33.3) |
| | ≤ Primary school | 47(40.1) |
| | ≤ Junior high school | 20(17.1) |
| | ≥ Senior high school | 11(9.4) |
| Employed status | Working | 81(69.2) |
| District | Jia-Sian | 43(36.8) |
|  | Shan-Lin | 74(63.2) |
| Distance to nearest clinic (km) | Mean ± SD | 4.26±3.71 |
| Distance to nearest hospital (km) | Mean ± SD | 21.68±10.90 |
| Current smoking | Yes | 24(20.5) |
| Current drinking | Yes | 21(17.9) |
| Current betelnut using | Yes | 9(7.7) |
| Past history |  |  |
| Cardiovascular diseases | Yes | 32(27.4) |
| Hypertension | Yes | 89(76.1) |
| Chronic hepatitis | Yes | 11(9.4) |
| Stroke | Yes | 9(7.7) |
| Chronic kidney disease | Yes | 18(15.4) |
| Malignancy | Yes | 4(3.4) |
| Surgical intervention | Yes | 60(51.3) |
| Laboratory data |  |  |
| Fasting plasma glucose (mg/dL) | Mean ± SD | 150.67±50.77 |
| | $\leq 130$ | 45(38.5) |
| | $>130$ | 58(49.6) |
| HbA1c (%) | Mean ± SD | 8.17±1.92 |
| | $< 7$ | 36(30.8) |
| | $\geq 7$ | 81(69.2) |
| Total cholesterol (mg/dL) | Mean ± SD | 200.45±49.02 |
| | $< 160$ | 24(20.5) |
| | $\geq 160$ | 86(73.5) |
| LDL-C (mg/dL) | Mean ± SD | 107.12±38.56 |
| | $< 100$ | 41(35.0) |
| | $\geq 100$ | 57(48.7) |
| HDL-C (mg/dL) in total | Goal-achieved | 61(52.1) |
|  | Not goal-achieved | 38(32.5) |
| HDL-C (mg/dL) in men | Mean ± SD | 48.07±14.09 |
| | $\geq 40$ | 29(24.8) |
|  | < 40 | 14(12.0) |
| HDL-C (mg/dL) in women | Mean ± SD | 54.93±13.77 |
| | $\geq 50$ | 38(32.5) |
|  | < 50 | 18(15.4) |
| TG (mg/dL) | Mean ± SD | 221.51±249.68 |
|  | < 150 | 48(41.0) |
| | $\geq 150$ | 59(50.4) |
| Proteinuria | No | 75(64.1) |
|  | Yes | 30(25.6) |

Abbreviations: HbA1c, glycohemoglobin; SD, standard deviation; LDL-C, low-density lipoprotein cholesterol; HDL, high-density lipoprotein cholesterol; TG, triglyceride

### Glycemic status and geographic accessibility

The overall mean HbA1c level among the subjects was 8.17% (±1.92), with approximately 70% of the T2DM patients in this study categorized within the “poor control” group (HbA1c ≥ 7%). The average distance from the patient’s residence to the nearest clinic was 4.26 kilometers (±3.71), and to the nearest hospital was 21.68 kilometers (±10.90). According to Spearman’s rank correlation test, as shown in Table 2, there was a modest correlation between the distance to both the hospital or clinic and the HbA1c levels, suggesting that greater geographic distances to medical facilities may be associated with poorer glycemic control.

**Table 2.** Result of Spearman’s Rank Correlation Analysis.

|  | Correlation coefficient (Rho) | *p*-value |
| --- | --- | --- |
| Distance to the nearest clinic | 0.252 | 0.006* |
| Distance to the nearest hospital | 0.241 | 0.009* |

\**p* < 0.05

### Glycemic control and categoric variables

As presented in Table 3, good glycemic control was significantly associated with several covariates, including employment status (*p* = 0.010), residential district (*p* = 0.010), control of fasting plasma glucose (*p* = 0.003), triglyceride levels (*p* = 0.049), and high-density lipoprotein cholesterol levels (*p* = 0.020). We further applied logistic regression to assess the influence of these factors on glycemic control, with the findings presented in Table 4. Both of univariate and multivariate analysis indicated that unemployment, residing in the Shan-Lin district, achieving target fasting plasma glucose levels, and maintaining optimal HDL-C levels were factors statistically associated with good glycemic control.

**Table 3.** Results of Chi-square test or Fisher’s exact test for glycemic control and categorical variables.

| Characteristics | Categories | HbA1c < 7% (n) | HbA1c ≥ 7% (n) | *p*-value |
| --- | --- | --- | --- | --- |
| General data |  |  |  |  |
| Age(years) | <65 | 15 | 48 | 0.078 |
|  | ≥65 | 21 | 33 |  |
| Sex | Male | 15 | 32 | 0.826 |
|  | Female | 21 | 49 |  |
| Education level | Unschool | 11 | 28 | 0.451 |
|  | ≤Primary school | 12 | 35 |  |
|  | ≤Junior high school | 8 | 12 |  |
|  | ≥Senior high school | 5 | 6 |  |
| Employed status | Not working | 17 | 19 | 0.010* |
|  | Working | 19 | 62 |  |
| Address district | Jia-Sian | 7 | 36 | 0.010* |
|  | Shan-Lin | 29 | 45 |  |
| Current smoking | No | 27 | 66 | 0.423 |
|  | Yes | 9 | 15 |  |
| Current drinking | No | 29 | 67 | 0.799 |
|  | Yes | 7 | 14 |  |
| Betelnut using | No | 32 | 76 | 0.454 |
|  | Yes | 4 | 5 |  |
| Past history |  |  |  |  |
| Cardiovascular diseases | No | 25 | 60 | 0.604 |
|  | Yes | 11 | 21 |  |
| Hypertension | No | 11 | 16 | 0.213 |
|  | Yes | 25 | 64 |  |
| Chronic hepatitis | No | 33 | 73 | 1.000 |
|  | Yes | 3 | 8 |  |
| Stroke | No | 32 | 76 | 0.454 |
|  | Yes | 4 | 5 |  |
| Chronic kidney disease | No | 27 | 72 | 0.055 |
|  | Yes | 9 | 9 |  |
| Malignancy | No | 36 | 77 | 0.310 |
|  | Yes | 0 | 4 |  |
| Surgical intervention | No | 16 | 40 | 0.580 |
|  | Yes | 20 | 40 |  |
| Laboratory data |  |  |  |  |
| Fasting plasma glucose (mg/dL) | ≤ 130 | 20 | 25 | 0.003* |
|  | > 130 | 10 | 48 |  |
| Total cholesterol (mg/dL) | < 160 | 5 | 19 | 0.447 |
|  | ≥ 160 | 27 | 59 |  |
| LDL-C (mg/dL) | < 100 | 13 | 28 | 0.727 |
|  | ≥ 100 | 20 | 37 |  |
| HDL-C (mg/dL) in total | Goal-achieved | 23 | 38 | 0.020* |
|  | Not goal-achieved | 6 | 32 |  |
| HDL-C (mg/dL) in men | ≥ 40 | 9 | 20 | 1.000 |
|  | < 40 | 5 | 9 |  |
| HDL-C (mg/dL) in women | ≥ 50 | 14 | 24 | 0.021* |
|  | < 50 | 1 | 17 |  |
| TG (mg/dL) | < 150 | 18 | 30 | 0.049* |
|  | ≥ 150 | 12 | 47 |  |
| Proteinuria | No | 25 | 50 | 0.315 |
|  | Yes | 7 | 23 |  |

Abbreviations: HbA1c, glycohemoglobin; SD, standard deviation; LDL-C, low-density lipoprotein cholesterol; HDL, high-density lipoprotein cholesterol; TG, triglyceride
\* $p < 0.05$

**Table 4.**
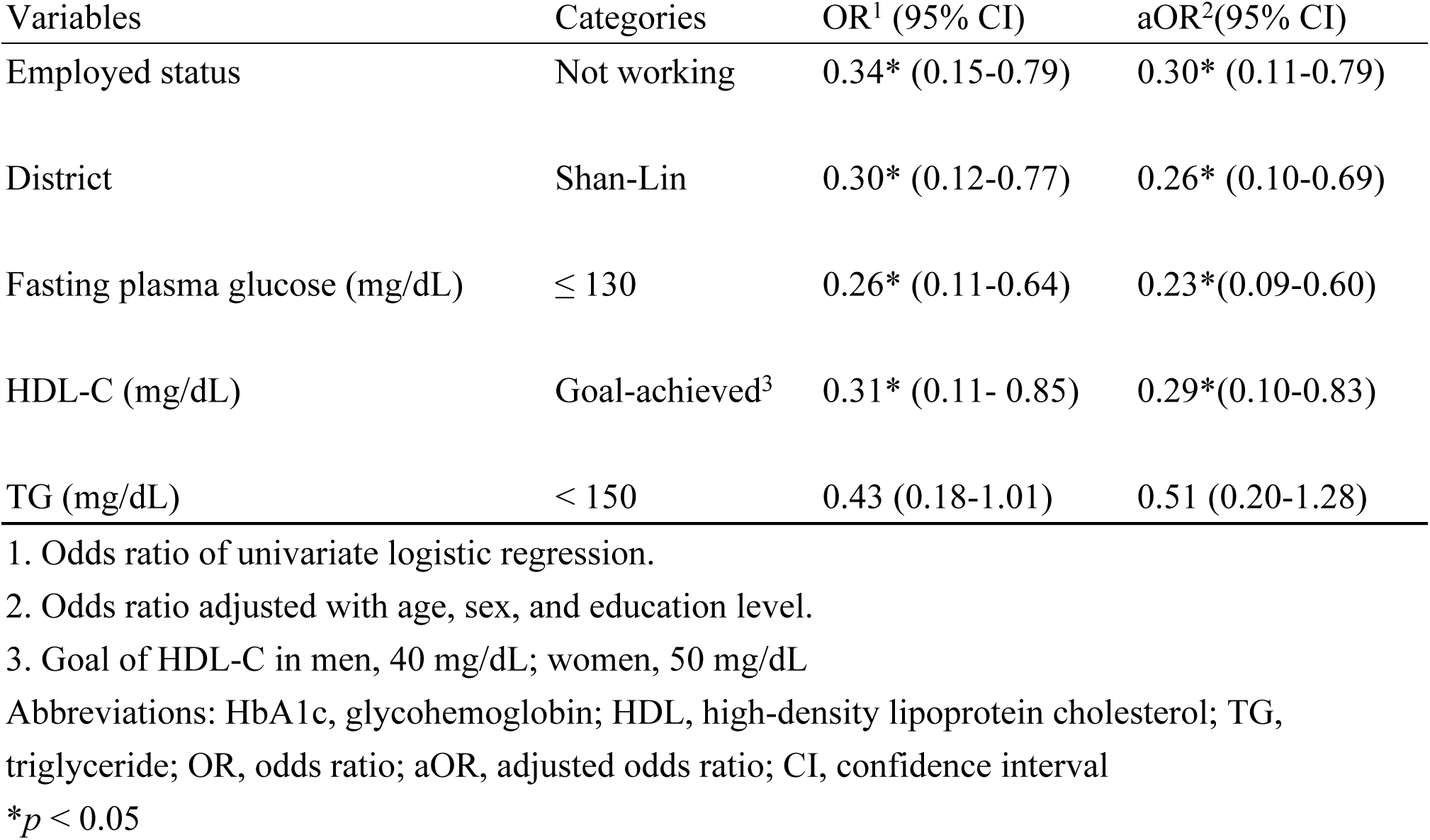
Logistic regression of variables associated with good control of HbA1c.

| Variables | Categories | OR<sup>1</sup> (95% CI) | aOR<sup>2</sup>(95% CI) |
| --- | --- | --- | --- |
| Employed status | Not working | 0.34* (0.15-0.79) | 0.30* (0.11-0.79) |
| District | Shan-Lin | 0.30* (0.12-0.77) | 0.26* (0.10-0.69) |
| Fasting plasma glucose (mg/dL) | ≤ 130 | 0.26* (0.11-0.64) | 0.23*(0.09-0.60) |
| HDL-C (mg/dL) | Goal-achieved<sup>3</sup> | 0.31* (0.11- 0.85) | 0.29*(0.10-0.83) |
| TG (mg/dL) | < 150 | 0.43 (0.18-1.01) | 0.51 (0.20-1.28) |

1. Odds ratio of univariate logistic regression.
2. Odds ratio adjusted with age, sex, and education level.
3. Goal of HDL-C in men, 40 mg/dL; women, 50 mg/dL
Abbreviations: HbA1c, glycohemoglobin; HDL, high-density lipoprotein cholesterol; TG, triglyceride; OR, odds ratio; aOR, adjusted odds ratio; CI, confidence interval
\* $p < 0.05$

## Discussion

The primary objective of this study is to uncover the relationship between geographic accessibility and long-term glycemic status among rural Taiwanese populations with type 2 diabetes mellitus. Our findings, as manifested in Table 2, reveal a modest but positive correlation between the distance from a patient’s residence to both primary (clinic) and secondary (hospital) healthcare facilities and HbA1c level. This extrapolates that rural T2DM patients residing further from medical units are more likely to experience chronic hyperglycemia. This observation may help explain the higher rates of diabetic complications and the risk of avoidable hospitalizations noted in previous Taiwanese rural studies[9–11]. The influence of geographical location on glycemic control corroborates findings from studies in other countries. In the United States, Strauss, K. et al. observed that rural diabetics with longer driving distances from home to primary care providers tended to exhibit poor glycemic control[16].

Similarly, in a Pennsylvanian study, Zgibor, J. C. et al. found that T2DM patients living within 10 miles of a medical center were 2.5 times more likely to have lower HbA1c levels between first and last office visits[19]. However, a Japanese rural study reported inconsistent spatial effects in different medical units of reference. According to R. Fukuoka et al., HbA1c level was found to be associated with distance to secondary medical facilities rather than primary ones[13]. In the author’s opinion, the discrepancy is possibly related to patients’ preference on specialized doctor in larger hospitals. Interestingly, in our study, distance to the nearest clinic and hospital share similar correlation coefficient (Rho: 0.252 versus 0.241), despite a notable difference in average distances (4.26 km versus 21.68 km). This indicates that clinics seem to play as a comparable role to hospitals which typically offer more advanced devices and a greater number of specialists in this study. This counter-intuitive finding could be attributed to Taiwan government’s promotion of diabetic care utilization in clinics. According to national statistics, diabetic treatment in outpatient care has been evenly distributed among clinics, hospitals, and medical centers since 2009[4]. Additionally, nationwide studies in Taiwan have indicated a reduction in the rural-urban disparity in diabetic care at primary health units[11, 20], reflecting effective public health strategies to enhance accessibility and care continuity for rural populations.

In addition to spatial factors, our study also examined social and biochemical determinants that may influence glycemic control. Notably, nearly half of the study’s participants were elderly, double the proportion of geriatric individuals in the global diabetic population[21]. Aging population rises a concern that healthcare providers in rural region need to pay more attention to geriatric complications. Furthermore, the majority of our subjects had an education level below junior high school. Low educational attainment has been linked to increased risks of developing conditions such as coronary heart disease, chronic kidney disease, and diabetic foot[22–24]. Comorbidities such as hypertension and cardiovascular diseases were prevalent among our subjects, aligning with findings from previous rural studies in Taiwan that reported a 12% higher risk of cardiovascular events compared to the urban diabetic population[5].

Our logistic regression analysis identified several variables associated with better glycemic control (HbA1c < 7 %). Not-working status seemed to be a protective factor against chronic glycemia. In rural settings, non-working individuals were often retirees or homemakers who may have more time to engage in health-promoting activities such as exercising, maintaining healthy diets, and participating in health education programs at medical facilities. Another key finding was that T2DM patients living in Shan-Lin district were more likely to experience good glycemic control.

Given that Shan-Lin district is closer to the hospital and downtown compared to Jia-Sian, this could suggest a link between proximity to healthcare resources and long-term glycemic control. However, further qualitative study is needed to explore other socio-economic factors that might influence this association. Additionally, achieving goals for fasting glucose and HDL-C were both positively correlated with controlled HbA1c levels. Previous studies have documented the close relationship between fasting glucose levels and HbA1c[25, 26]. Moreover, HDL dysfunction has been shown to adversely affect pancreatic β-cell function and insulin sensitivity in muscle tissues[27], which may exacerbate the progression of T2DM[28]. In rural diabetic care, healthcare providers often face challenges in regularly assessing patients’ HbA1c levels. Recognizing the factors discussed above can be beneficial in evaluating patients’ long-term glycemic status and tailoring appropriate interventions. This holistic approach is essential for improving diabetes management and outcomes in rural populations.

This study has several limitations that warrant consideration. Firstly, the relatively small sample size of T2DM patients may lead to wide confidence intervals of odds ratio in the logistic regression analysis. This limitation likely arises from the stringent inclusion criteria that subjects have HbA1c values measured within three months of their initial visit to the Mobile Health Care outpatient service. To more precisely determine the influence among factors on glycemic control, larger cohort studies are necessary. Secondly, some critical factors that may be related to glycemic control, such as body mass index, duration of T2DM, marital status, and income, were not available in this retrospective study. The absence of these variables could limit the comprehensiveness and accuracy of our findings. Thirdly, distance calculated by Google Maps from a patient’s address to the nearest medical unit may not perfectly reflect geographic accessibility. Patients may choose different routes or utilize various transportation to medical units, introducing variability that could reduce the precision of the estimates. However, given the paucity of medical facilities and inconvenient public transportation in the rural areas studied, assuming that the distance to the nearest clinic or hospital reasonably represents the geographic burden faced by patients in accessing diabetic care remains a practical and justified approach.

## Conclusion

This study highlights the influence of geographic accessibility and socio-economic factors on long-term glycemic control in rural diabetic patients in Taiwan. By demonstrating a positive correlation between distance to medical facilities and HbA1c levels, our findings suggest that patients residing further from healthcare resources tend to experience poorer glycemic control. This geographical challenge underscores the necessity for policy makers to consider distance as a significant factor in rural diabetes management. Moreover, our research identifies several indicators such as control of fasting glucose and lipid profiles, employment status, and address district that can help rural physicians rapidly assess potential long-term glycemic control upon a patient’s initial presentation.

In conclusion, this study offers valuable insights of glycemic control for rural healthcare providers and policy makers, underscoring the need for targeted interventions to improve access to diabetic care in rural areas, such as mobile health services and community-based health education programs. This research also calls for larger-scale studies to further investigate the causal relationships and to explore additional factors affecting glycemic control in rural diabetic population, which could inform policy and healthcare strategies tailored to these communities.

## Data Availability

Main data are within the manuscript. Other precise individual data are available from contact email of corresponding author.

## Acknowledgements

The authors would like to express gratitude to medical team of Mobile Health Care for collecting subjects’ profiles and appreciate the Biostatistics Center, Kaohsiung Chang Gung Memorial Hospital for statistics work.

## Reference

1. International Diabetes Federation.IDF Diabetes Atlas, 10th edn. Brussels, Belgium: 2021. Available from: https://www.diabetesatlas.org.

2. Sheen YJ, Hsu CC, Jiang YD, Huang CN, Liu JS, Sheu WH. Trends in prevalence and incidence of diabetes mellitus from 2005 to 2014 in Taiwan. J Formos Med Assoc. 2019;118 Suppl 2:S66–s73. Epub 20190709. doi: 10.1016/j.jfma.2019.06.016. PubMed PMID: 31300322.

3. Bommer C, Heesemann E, Sagalova V, Manne-Goehler J, Atun R, Bärnighausen T, et al. The global economic burden of diabetes in adults aged 20-79 years: a cost-of-illness study. Lancet Diabetes Endocrinol. 2017;5(6):423–30. Epub 20170426. doi: 10.1016/s2213-8587(17)30097-9. PubMed PMID: 28456416.

4. Wang CY, Wu YL, Sheu WH, Tu ST, Hsu CC, Tai TY. Accountability and utilization of diabetes care from 2005 to 2014 in Taiwan. J Formos Med Assoc. 2019;118 Suppl 2:S111–S21. Epub 20191004. doi: 10.1016/j.jfma.2019.08.010. PubMed PMID: 31590971.

5. Tai SY, He JS, Kuo CT, Kawachi I. Urban-Rural Disparities in the Incidence of Diabetes-Related Complications in Taiwan: A Propensity Score Matching Analysis. J Clin Med. 2020;9(9). Epub 20200918. doi: 10.3390/jcm9093012. PubMed PMID: 32962006; PubMed Central PMCID: PMCPMC7565280.

6. Chatterjee S, Davies MJ, Heller S, Speight J, Snoek FJ, Khunti K. Diabetes structured self-management education programmes: a narrative review and current innovations. Lancet Diabetes Endocrinol. 2018;6(2):130–42. Epub 20170929. doi: 10.1016/s2213-8587(17)30239-5. PubMed PMID: 28970034.

7. Dugani SB, Mielke MM, Vella A. Burden and management of type 2 diabetes in rural United States. Diabetes Metab Res Rev. 2021;37(5):e3410. Epub 20201005. doi: 10.1002/dmrr.3410. PubMed PMID: 33021052; PubMed Central PMCID: PMCPMC7990742.

8. Wang Q, Zhang X, Fang L, Guan Q, Guan L, Li Q. Prevalence, awareness, treatment and control of diabetes mellitus among middle-aged and elderly people in a rural Chinese population: A cross-sectional study. PLoS One. 2018;13(6):e0198343. Epub 20180601. doi: 10.1371/journal.pone.0198343. PubMed PMID: 29856828; PubMed Central PMCID: PMCPMC5983453.

9. Li CH, Li CC, Lu CL, Wu JS, Ku LE, Li CY. Urban-rural disparity in lower extremities amputation in patients with diabetes after nearly two decades of universal health Insurance in Taiwan. BMC Public Health. 2020;20(1):212. Epub 20200211. doi: 10.1186/s12889-020-8335-3. PubMed PMID: 32046698; PubMed Central PMCID: PMCPMC7014711.

10. Lee CM, Chang CC, Pan MY, Chang CF, Chen MY. Insufficient early detection of peripheral neurovasculopathy and associated factors in rural diabetes residents of Taiwan: a cross-sectional study. BMC Endocr Disord. 2014;14:89. Epub 20141124. doi: 10.1186/1472-6823-14-89. PubMed PMID: 25421066; PubMed Central PMCID: PMCPMC4255455.

11. Chen CC, Chen LW, Cheng SH. Rural-urban differences in receiving guideline-recommended diabetes care and experiencing avoidable hospitalizations under a universal coverage health system: evidence from the past decade. Public Health. 2017;151:13–22. Epub 20170708. doi: 10.1016/j.puhe.2017.06.009. PubMed PMID: 28697373.

12. Biswas RK, Kabir E. Influence of distance between residence and health facilities on non-communicable diseases: An assessment over hypertension and diabetes in Bangladesh. PLoS One. 2017;12(5):e0177027. Epub 20170518. doi: 10.1371/journal.pone.0177027. PubMed PMID: 28545074; PubMed Central PMCID: PMCPMC5436657.

13. Fukuoka R, Takeda M, Abe T, Yamasaki M, Kimura S, Okuyama K, et al. Inconvenience of Living Place Affects Individual HbA1c Level in a Rural Area in Japan: Shimane CoHRE Study. Int J Environ Res Public Health. 2021;18(3). Epub 20210128. doi: 10.3390/ijerph18031147. PubMed PMID: 33525428; PubMed Central PMCID: PMCPMC7908499.

14. LePage AK, Wise JB, Bell JJ, Tumin D, Smith AW. Distance from the endocrinology clinic and diabetes control in a rural pediatric population. J Pediatr Endocrinol Metab. 2021;34(2):187–93. Epub 20201012. doi: 10.1515/jpem-2020-0332. PubMed PMID: 33544546.

15. Nouhjah S, Shahbazian H, Jahanfar S, Shahbazian N. The effect of distance on the adherence to postpartum follow-up in women with gestational diabetes. Environ Sci Pollut Res Int. 2021;28(46):65428–34. Epub 20210728. doi: 10.1007/s11356-021-15472-3. PubMed PMID: 34318425.

16. Strauss K, MacLean C, Troy A, Littenberg B. Driving distance as a barrier to glycemic control in diabetes. J Gen Intern Med. 2006;21(4):378–80. doi: 10.1111/j.1525-1497.2006.00386.x. PubMed PMID: 16686817; PubMed Central PMCID: PMCPMC1484707.

17. ElSayed NA, Aleppo G, Aroda VR, Bannuru RR, Brown FM, Bruemmer D, et al. 2. Classification and Diagnosis of Diabetes: Standards of Care in Diabetes-2023. Diabetes Care. 2023;46(Supplement_1):S19-s40. doi: 10.2337/dc23-S002. PubMed PMID: 36507649.

18. DAROC Clinical Practice Guidelines for Type 2 Diabetes Care-2022, Taiwan, Diabetes Association of the R.O.C., 2022.

19. Zgibor JC, Gieraltowski LB, Talbott EO, Fabio A, Sharma RK, Hassan K. The association between driving distance and glycemic control in rural areas. J Diabetes Sci Technol. 2011;5(3):494–500. Epub 20110501. doi: 10.1177/193229681100500304. PubMed PMID: 21722565; PubMed Central PMCID: PMCPMC3192616.

20. Cheng BR, Chang HT, Lin MH, Chen TJ, Chou LF, Hwang SJ. Rural-urban disparities in family physician practice patterns: A nationwide survey in Taiwan. Int J Health Plann Manage. 2019;34(1):e464–e73. Epub 20180920. doi: 10.1002/hpm.2662. PubMed PMID: 30238506.

21. International Diabetes Federation (IDF) (2015) Diabetes Atlas. 7th Edition, International Diabetes Federation, Brussels, Belgium. Available from: http://www.diabetesatlas.org.

22. Pourkazemi A, Ghanbari A, Khojamli M, Balo H, Hemmati H, Jafaryparvar Z, et al. Diabetic foot care: knowledge and practice. BMC Endocr Disord. 2020;20(1):40. Epub 20200320. doi: 10.1186/s12902-020-0512-y. PubMed PMID: 32192488; PubMed Central PMCID: PMCPMC7083045.

23. Mizouri R, Belhadj M, Hasni Y, Maaroufi A, Mahjoub F, Jamoussi H. Relationship between level of education and podiatry risk in diabetic patients. Tunis Med. 2021;99(2):277–84. PubMed PMID: 33899199; PubMed Central PMCID: PMCPMC8636960.

24. Slåtsve KB, Claudi T, Lappegård KT, Jenum AK, Larsen M, Nøkleby K, et al. Level of education is associated with coronary heart disease and chronic kidney disease in individuals with type 2 diabetes: a population-based study. BMJ Open Diabetes Res Care. 2022;10(5). doi: 10.1136/bmjdrc-2022-002867. PubMed PMID: 36171015; PubMed Central PMCID: PMCPMC9528574.

25. Ghazanfari Z, Haghdoost AA, Alizadeh SM, Atapour J, Zolala F. A Comparison of HbA1c and Fasting Blood Sugar Tests in General Population. Int J Prev Med. 2010;1(3):187–94. PubMed PMID: 21566790; PubMed Central PMCID: PMCPMC3075530.

26. Mayega RW, Guwatudde D, Makumbi FE, Nakwagala FN, Peterson S, Tomson G, et al. Comparison of fasting plasma glucose and haemoglobin A1c point-of-care tests in screening for diabetes and abnormal glucose regulation in a rural low income setting. Diabetes Res Clin Pract. 2014;104(1):112–20. Epub 20140103. doi: 10.1016/j.diabres.2013.12.030. PubMed PMID: 24456993.

27. Rotllan N, Julve J, Escolà-Gil JC. Type 2 Diabetes and HDL dysfunction: a key contributor to Glycemic Control. Curr Med Chem. 2023. Epub 20230201. doi: 10.2174/0929867330666230201124125. PubMed PMID: 36722477.

28. Waldman B, Jenkins AJ, Sullivan D, Ng MKC, Keech AC. HDL as a Target for Glycemic Control. Curr Drug Targets. 2017;18(6):651–73. doi: 10.2174/1389450116666150727115544. PubMed PMID: 26212264.

